# Characterization of *Mycobacterium tuberculosis*-specific Th22 cells and the effect of tuberculosis disease and HIV co-infection

**DOI:** 10.1101/2020.10.22.20217521

**Authors:** Mohau S. Makatsa, F. Millicent A. Omondi, Rubina Bunjun, Robert J. Wilkinson, Catherine Riou, Wendy A. Burgers

**Author notes:** **Corresponding author:** Wendy Burgers, Institute of Infectious Disease and Molecular Medicine, University of Cape Town, Falmouth Building, Room 3.36.1, Observatory 7925, South Africa. C.R. and W.A.B. contributed equally to this work.

## Abstract

The development of a highly effective tuberculosis (TB) vaccine is likely dependent on our understanding of what constitutes a protective immune response to TB. Accumulating evidence suggests that CD4+ T cells producing IL-22, a distinct subset termed ‘Th22’ cells, may contribute to protective immunity to TB. Thus, we characterized *Mycobacterium tuberculosis* (Mtb)-specific Th22 (and Th1 and Th17) cells in 72 individuals with latent tuberculosis infection (LTBI) or TB disease, with and without human immunodeficiency virus (HIV)-1 infection. We investigated the functional properties (IFN-γ, IL-22 and IL-17 production), memory differentiation (CD45RA, CD27 and CCR7) and activation profile (HLA-DR) of Mtb-specific CD4+ T cells. In HIV-uninfected individuals with LTBI, we detected abundant IFN-γ producing CD4+ T cells (median: 0.93%) and IL-22-producing CD4+ T cells (median: 0.46%) in response to Mtb. The frequency of IL-17 producing CD4+ T cells was much lower, at a median of 0.06%. Consistent with previous studies, IL-22 was produced by a distinct subset of CD4+ T cells and not co-expressed with IL-17. Mtb-specific IL-22 responses were markedly reduced (median: 0.08%) in individuals with TB disease and HIV co-infection compared to IFN-γ responses. Mtb-specific Th22 cells exhibited a distinct memory and activation phenotype compared to Th1 and Th17 cells. Furthermore, Mtb-specific IL-22 was produced by conventional CD4+ T cells that required T cell receptor (TCR) engagement. In conclusion, we confirm that Th22 cells contribute substantially to the immune response to TB. Depletion of Mtb-specific Th22 cells during HIV co-infection may contribute to increased risk of TB disease.

## INTRODUCTION

Tuberculosis (TB) is the leading cause of death from an infectious disease worldwide, with 10 million TB cases per year and 1.6 million deaths in 2018 (1). South Africa is one of the eight countries that together account for two thirds of all TB cases globally (1). Moreover, approximately one quarter of the world’s population is latently infected with *Mycobacterium tuberculosis* (Mtb) (1). In immunocompetent individuals, the risk of progression from infection to active TB disease is 2–10% in a lifetime, illustrating that the human immune system can control Mtb in most cases (2). Bacillus Calmette-Guérin (BCG) is the only licensed TB vaccine and protects against disseminated TB in children but provides variable protection against highly prevalent pulmonary tuberculosis (PTB) in adults (3, 4). Therefore, there is an urgent need for new and effective TB vaccines, and recent progress in clinical and pre-clinical trials have delivered promising results (5–7).

A better understanding of the immune responses required to control Mtb will aid the development of improved vaccines against TB (8). It is well established that CD4+ T cells, particularly Th1 cells producing the cytokines IFN-γ and TNF-α, are critical for immunity against Mtb (8, 9). However, Th1 immunity alone is not sufficient, as IFN-γ has been reported to be a poor correlate of BCG vaccination-induced protection against TB in mice (10, 11). Furthermore, IFN-γ-independent mechanisms of CD4+ T cell mediated control of Mtb infection have been documented (12–14). Thus, other CD4+ T helper subsets beyond Th1 cells may be essential for protection against TB.

There is growing interest in the cytokine IL-22 and its role in TB immunity. IL-22 belongs to the IL-10 family of cytokines and its receptor is composed of two heterodimeric subunits, IL-22R1 and IL-10R2 (15). IL-22 mainly targets non-hematopoietic cells, namely epithelial cells, and fibroblasts in tissues (15), but expression of the IL-22 receptor has also been reported on macrophages (16, 17). IL-22 promotes tissue proliferation, regeneration, and healing (15-20). It induces the production of antimicrobial peptides and proteins such as β-defensins, the S100 family of peptides, Reg3, calprotectin and calgranulin A (18–20). Furthermore, IL-22 signaling regulates chemokine expression to orchestrate the recruitment of immune cell subsets to sites of infection (20, 21). During infection with Mtb, IL-22 was initially reported to be dispensable for Mtb control in mouse models (22, 23). Recently, however, a protective role for IL-22 in TB immunity was described in a murine model, where IL-22 deficient mice displayed greater bacterial burdens after aerosol infection with a virulent clinical strain of Mtb, HN878 (17). In humans, soluble IL-22 has been detected at sites of extra-pulmonary tuberculosis (24), and a higher concentration of IL-22 was observed in bronchoalveolar lavage fluid (BALF) of individuals with active TB compared to healthy donors (25). Moreover, IL-22 has been shown to inhibit intracellular *M. tuberculosis* growth in macrophages (16), and a polymorphism in the IL-22 promoter has been linked to increased TB risk (26).

IL-22 is produced by a variety of cells, including T cells (Th17, Th1 and γδ T cells) and innate cells (innate lymphoid cells (ILCs) and NK cells) (16, 27, 28). In humans, IL-22 is mainly produced by a distinct subset of CD4+ T cells, named Th22 cells (29, 30). Our laboratory recently showed that IL-22-producing CD4+ T cells contribute to the mycobacterial response during latent TB infection (LTBI) (31). These mycobacteria-specific Th22 cells were depleted during HIV infection to a similar extent as Th1 cells, emphasizing their potential importance in protective immunity to TB in the context of HIV. In the present study, we further characterized Mtb-specific Th22 cells during TB disease and HIV co-infection. This study investigated the contribution of Th22 cells to TB immunity, characterized Th22 cells further to gain insights into their function, and examine the impact of TB disease and HIV infection on Th22 cells, in comparison to Th1 and Th17 cells. Our findings confirm that IL-22 is produced by CD4+ T cells in response to Mtb antigens, and these Th22 cells contribute substantially to Mtb immune responses. Moreover, Mtb-specific Th22 cells were severely diminished in patients with both HIV infection and TB disease. Additionally, we demonstrate for the first time that IL-22 production is dependent on TCR engagement but may require alternative co-stimulatory molecules or antigen presenting cells.

## MATERIAL AND METHODS

### Study participants

Blood samples were collected from 72 individuals recruited from the Ubuntu Clinic, Khayelitsha, Cape Town, South Africa. Participants were classified into four groups according to their HIV-1 infection status and whether they had latent TB infection (LTBI) or active TB disease (aTB): HIV-/ LTBI (*n* = 19), HIV+/LTBI (*n* = 18), HIV-/aTB (*n* = 19), and HIV+/aTB (*n* = 16). The clinical characteristics for each group are presented in **Table 1**. LTBI was diagnosed based on a positive IFN-γ release assay (QuantiFERON®-TB Gold In-Tube), no symptoms of aTB and no detection of Mtb in sputum by GeneXpert. Diagnosis of aTB was based on clinical symptoms and positive sputum test (GeneXpert). All HIV-infected individuals were antiretroviral treatment (ART) naïve and all TB cases were drug sensitive and TB treatment-naïve at the time of enrolment. In addition to the patient cohort, healthy adult donors were recruited from the University of Cape Town for selected experiments. All participants gave written, informed consent. These studies were approved by the University of Cape Town, Faculty of Health Sciences Human Research Ethics Committee (HREC 279/2012 and HREC 896/2014).

**Table 1:** Clinical characteristics of the four study groups.

|  | LTBI |  | Active TB |  |
| --- | --- | --- | --- | --- |
|  | HIV– | HIV+ | HIV– | HIV+ |
| *n* | 19 | 18 | 19 | 16 |
| CD4 count (cells/mm<sup>3</sup>) <sup>a</sup> | 923<br>[687-1051] | 547<br>[463-626] | 648<br>[593-778] | 153<br>[75-207] |
| Log10 HIV viral load<br>(RNA copies/ml) <sup>a</sup> | n.a | 4.61<br>[3.72-4.97] | n.a | 5.06<br>[4.68-5.54] |
| Age <sup>a</sup> | 25 [19-31] | 33 [29-38] | 31 [26-40] | 34 [29-46] |
| Sex (F/M) | 9/10 | 16/2 | 2/17 | 9/7 |

<sup>a</sup> medians and [interquartile ranges]. F: female; M: male, LTBI: latent tuberculosis infection. n.a: not applicable.

### Blood collection and whole blood stimulation

Blood was collected in sodium heparin tubes and processed within 4 hours of collection. Whole blood assays were performed according the protocol described previously (32). Briefly, 0.5 ml of heparinized whole blood was stimulated with Mtb cell lysate (strain H37Rv, 10 μg/ml, BEI Resources), a Mtb-specific peptide pool consisting of 17 and 16 peptides covering the entire ESAT-6 (early secretory antigenic target-6) and CFP-10 (culture filtrate protein-10), respectively (4 μg/ml), bacillus Calmette-Guerin (BCG) (MOI of 4, *Mycobacterium bovis* Danish strain 1331, SSI) or gamma-irradiated Mtb whole cells (Strain H37Rv, 600 μg/ml, BEI Resources) at 37°C for a total of 12 h in the presence of the co-stimulatory antibodies anti-CD28 and anti-CD49d (1 μg /ml each; BD Biosciences). Unstimulated cells were incubated with co-stimulatory antibodies only. After 7 h, Brefeldin A (10 µg/ml; Sigma-Aldrich) was added. Red blood cells were lysed with Alternative Lysing Solution (ALS: 150 mM NH4Cl, 10 mM KHCO_3_, 1 mM Na_4_EDTA), and the cell pellet was subsequently stained with LIVE/DEAD™ Fixable Violet Stain (Molecular Probes) or LIVE/DEAD Fixable Near-IR Stain (Invitrogen). Cells were then fixed using FACS lysing solution (BD Biosciences) and cryopreserved in FCS containing 10% dimethyl sulfoxide (DMSO) for batch staining. For lymphocyte-specific protein tyrosine kinase (Lck) inhibition, whole blood from healthy individuals was incubated with increasing concentrations (1 μM to 60 μM) of Lck inhibitor (7-cyclopentyl-5-(4-phenoxyphenyl)-7H-pyrrolo[2,3-d] pyrimidin-4-ylamine, Sigma) for 30 min prior to cell stimulation.

### Intracellular Cytokine Staining and Flow Cytometry

Cryopreserved cells were thawed, washed and then permeabilized with Perm/Wash buffer (BD Biosciences). Cells were incubated at 4°C for 1 h with the following antibodies: CD19 Pacific Blue (6D5; Biolegend), CD14 Pacific Blue (M5E2; Biolegend), CD3 BV650 (OKT3; BD Biosciences), CD4 PerCP-Cy5.5 (L200; BD Biosciences), CD45RA BV570 (HI100; Biolegend), CD27 PE-Cy5 (1A4CD27; Beckman Coulter), CCR7 PECF594 (3D12; BD Biosciences), HLA-DR APC-Cy7 (L243; BD Biosciences), IFN-γ Alexa700 (B27; BD Biosciences), IL-17 Alexa 647 (N49-653; BD Biosciences), IL-22 PE (22URTI; e-Bioscience). In order to investigate which T cells produce IL-22 and IFN-γ, the following antibodies were used for surface and intracellular staining: CD3 BV650 (OKT3; BD Biosciences), CD4 ECD (T4; Beckman Coulter), CD8 QD705 (3B5; Life Technologies), CD56 PE-Cy7 (NCAM 16.2; BD Biosciences), Pan γδ PE (IMMU510; Beckman Coulter), CD161 Alexa 647 (HP-3G10; eBioscience), Vα7.2 BV510 (3C10; Biolegend), IFN-γ Alexa700 (B27; BD Biosciences), IL-22 BV421 (22URTI; eBioscience). Stained cells were acquired on a BD Fortessa and analyzed using FlowJo (v10, TreeStar). A positive cytokine response was defined as at least twice the background cytokine response from unstimulated cells. To define the phenotype of cytokine-producing cells, a cutoff of 20 cytokine events was used. The gating strategy applied is presented in **Supplemental Figure S1**.

### Determining the TCR Vβ repertoire of cytokine+ cells

The IOTest® Beta Mark TCR Vβ Repertoire Kit (Beckman Coulter) was used to determine the TCR Vβ repertoire of Mtb-specific IFN-γ or IL-22-producing CD4+ T cells. Briefly, cells from Mtb lysate-stimulated whole blood were stained with CD3 BV650 (OKT3; BD Biosciences), CD4 PerCP-Cy5.5 (L200; BD Biosciences), IFN-γ Alexa700 (B27; BD Biosciences), IL-22 APC (22JOP; eBioscience) and each of the eight vials containing mixtures of conjugated TCR Vβ antibodies corresponding to 24 different specificities.

### Statistical analysis

All statistical tests were performed using Prism (v6; GraphPad). Nonparametric tests were used for all comparisons. The Kruskal-Wallis test with Dunn’s Multiple Comparison test was used for multiple comparisons and the Mann–Whitney U test and Wilcoxon matched pairs test were used for unmatched and paired samples, respectively. A p value of < 0.05 was considered statistically significant.

## RESULTS

### Characterization of CD4 Th22 cells in the immune response to Mtb in HIV-/LTBI individuals

A subset of CD4+ T cells that produces the cytokine IL-22 in response to mycobacterial antigen stimulation has been described (29-31). We sought to further characterize Mtb-specific IL-22 production and define the relative contribution of IL-22 compared to more frequently measured responses, namely IFN-γ and IL-17. We first compared the magnitude of Mtb-specific IL-22-producing CD4+ T cells to IFN-γ and IL-17 responses, in whole blood from healthy individuals with latent Mtb infection (n = 19; **Figure 1A)**. Mtb-specific CD4+ T cell responses (producing any of the measured cytokines) were detected in all individuals (median: 1.34%, interquartile range (IQR): 0.97-2.52). The highest frequency was observed for IFN-γ responses (median: 0.93%, IQR: 0.40-1.60). Although lower, the magnitude of IL-22+ Mtb-specific CD4+ T cells was not significantly different from the IFN-γ response (median: 0.46%, IQR: 0.22-0.96). In contrast, IL-17 producing CD4+ T cells were detectable at much lower frequencies (median: 0.06%, IQR: 0.04-0.11), ∼16 to ∼8 fold lower than IFN-γ (p < 0.0001) and IL-22 (p = 0.0002) responses, respectively (**Figure 1B**). Using a Boolean gating strategy, we next assessed all possible cytokine combinations to determine the co-expression profile of IFN-γ, IL-22 and IL-17 in Mtb-specific CD4+ T cells. **Figure 1C** shows that the single IFN-γ response accounted for a median of 60% of the total Mtb response, and single IL-22-producing cells contributed a median of 37% to Mtb response. Cytokine co-expression was marginal, with only a small proportion of IFN-γ/ IL-22 co-expressing cells observed (median: 3.6%, IQR: 2-6). Only three out of 19 individuals produced IL-17 alone that contributed more than 5% to the total Mtb response (**Figure 1C**).

**Figure 1:**
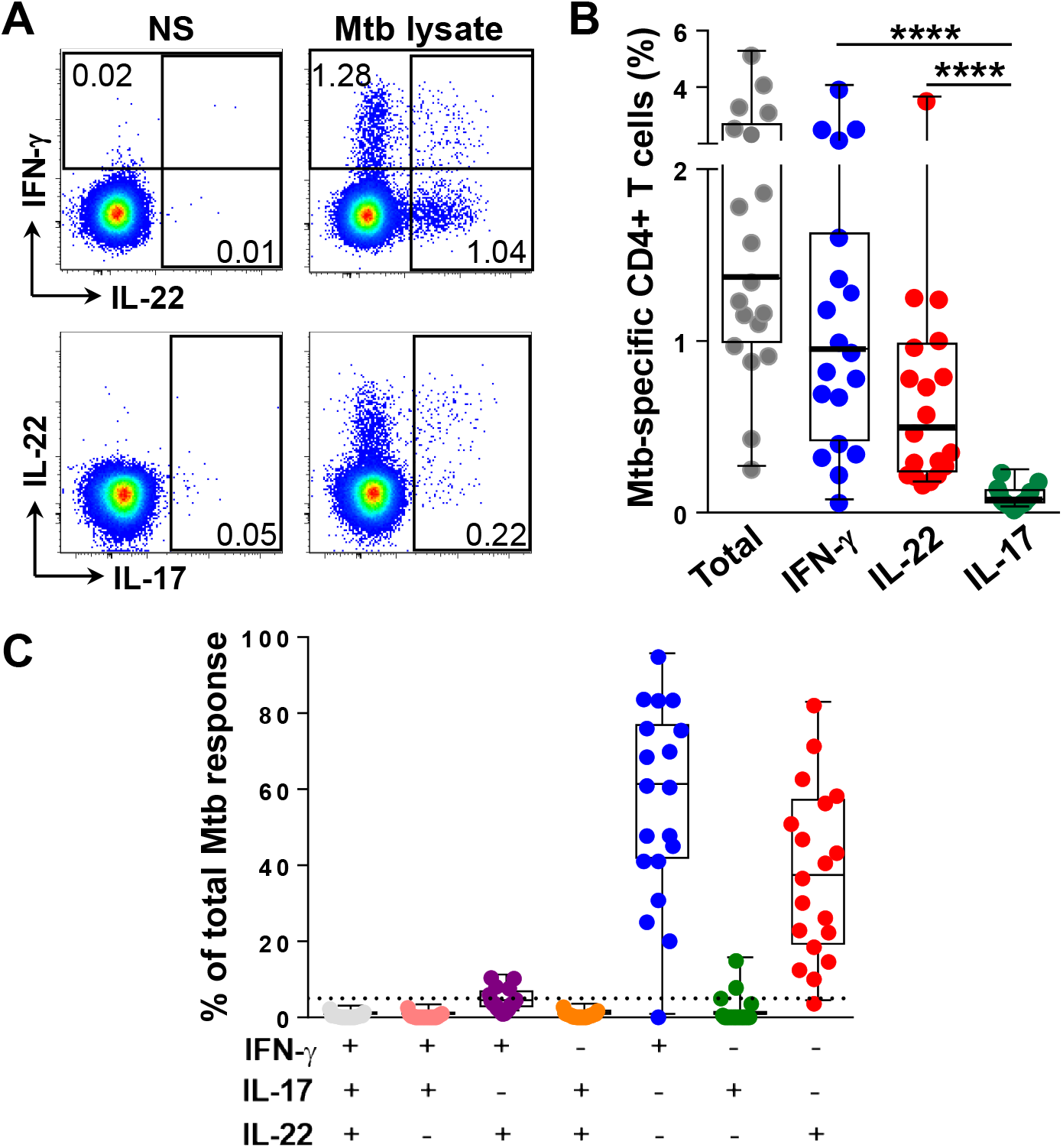
Contribution of IFN-γ, IL-22 and IL-17 to Mtb-specific CD4+ T cell responses in individuals with latent TB infection (LTBI). **(A)** Representative flow cytometry plots of IFN-γ, IL-22 and IL-17 production after stimulation with Mtb lysate. NS: no stimulation. **(B)** Summary graph of the frequency of cytokine responses. Statistical comparisons were performed using a one-way ANOVA Friedman test. \*\*\*\**p* < 0.0001. **(C)** Proportion of different combinations of IFN-γ, IL-22 and IL-17 in response to Mtb stimulation. Medians and interquartile ranges are depicted.

We next sought to determine whether similar cytokine profiles were generated in response to different types of Mtb antigens, and thus compared blood stimulated with Mtb lysate to gamma-whole cell Mtb (gamma-irradiated) and live mycobacteria (*M. bovis* BCG), as well as Mtb peptides, using an ESAT-6 and CFP-10 peptide pool. No differences in the magnitude of IL-22, IFN-γ or IL-17 responses were observed between the different “complex” mycobacterial antigens tested (*i*.*e*. Mtb cell lysate, gamma-irradiated Mtb and BCG), using 5 donors with LTBI (**Supplemental Figure S2A&B)**. This indicates that lysed, live or dead mycobacteria can detect IL-22 responses. In contrast, IL-22 production was barely detectable in response to Mtb peptide stimulation in 11 donors with LTBI (**Supplemental Figure 2C)**. These data raised two questions, namely (i) whether IL-22 production originates from unconventional T cells; or (ii) is the result of T cell receptor (TCR)-independent bystander activation, rather than being induced by CD4+ T cells upon engagement of cognate antigen presented to the TCR (33). A variety of immune cells have the ability to produce IL-22 (34, 35). Thus to address the first question, we investigated IL-22 and IFN-γ production from a range of T cell subsets in whole blood, including mucosal-associated invariant T (MAIT) cells, γδ T cells, Natural Killer T (NKT) cells, CD4+ T cells and CD8+ T cells. Samples were gated on live lymphocytes that were CD3+, and then defined as NKT (CD56+), γδ (CD56-γδTCR+), MAIT (CD56-γδTCR-Vα7.2+CD161+), CD4 (NKT-γδTCR-MAIT-CD8-CD4+) and CD8 (NKT-γδTCR-MAIT-CD4-CD8+) T cells (**Figure 2A**). In 10 healthy donors tested for cytokine responses, all cell subsets of interest produced IFN-γ when stimulated with Mtb lysate. In contrast, in 7/10 donors, IL-22 was exclusively produced by CD4+ T cells, with only 3 donors demonstrating a minor contribution of either MAIT or γδ T cells to the total IL-22 response from CD3+ cells (**Figure 2B**). Of note, the majority of MAIT and γδ T cells were CD4- (data not shown), indicating that our gating strategy used in the previous experiments was not likely to have included IL-22 from these T cell sources. Thus, conventional CD3+CD4+ T cells appear to be the major source of IL-22 in response to Mtb antigens.

**Figure 2:**
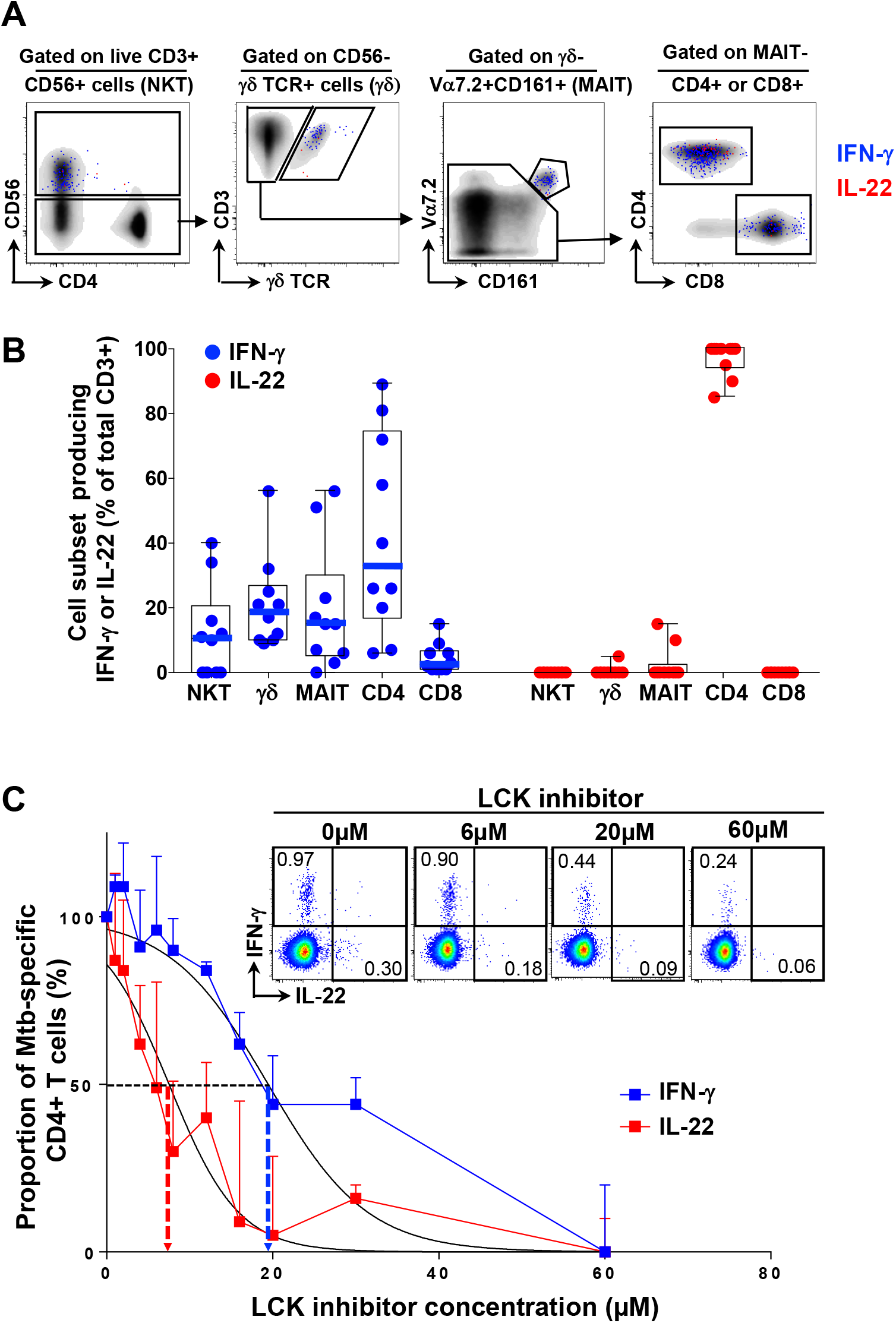
IL-22 responses from different cell populations and the impact of TCR blocking on IFN-γ and IL-22 production. (A) Representative flow cytometry plots showing the gating strategy for NKT, γδ, MAIT, CD4+ and CD8+ T cells. The overlaid dots indicate IFN-γ (blue) or IL-22 (red) production from live CD3+ lymphocytes in response to Mtb lysate. **(B)** The frequencies of each subset are indicated, as a percentage of total live CD3+ cells. (B) IFN-γ (blue) and IL-22 (red) responses to Mtb lysate from NKT, γδ, MAIT, CD4+ and CD8+ T cells in healthy donors (n=10). Each dot represents one individual. **(C)** Representative flow cytometry plots showing Mtb-specific IFN-γ and IL-22 responses in the presence of different concentrations of Lck inhibitor 7-Cyclopentyl-5-(4-phenoxyphenyl)-7H-pyrrolo[2,3-d] pyrimidin-4-ylamine and summary graph (n = 5). Medians and interquartile ranges are depicted. Vertical arrows show ED_50_. A nonlinear regression curve fit was used.

In order to determine whether IL-22 production was Mtb-specific (*i*.*e*. stimulated via recognition of cognate antigen by the TCR) or due to a bystander effect of cytokine activation of non-specific cells, we inhibited the TCR pathway by blocking Lck (a tyrosine kinase critical for early propagation of TCR signalling), using increasing concentrations of the Lck inhibitor 7-cyclopentyl-5-(4-phenoxyphenyl)-7H-pyrrolo[2,3-d] pyrimidin-4-ylamine. Using whole blood from five donors with LTBI, we found that as for IFN-γ, IL-22 production was suppressed upon Lck inhibition in a dose-dependent manner. Interestingly, the ED_50_ (efficient dose causing 50% of maximum effect) for IL-22 inhibition was 2.6 times lower compared to that of IFN-γ inhibition (7.5 µM and 19.5 µM, respectively; **Figure 2C**). These data demonstrate that IL-22 production from CD4+ T cells is TCR-dependent.

Lastly, we compared the TCRβ repertoire of IL-22 and IFN-γ-producing CD4+ T cells in blood from five donors with LTBI, using a commercial typing test. Of note, while the kit detects 24 out of 64 known TCR vβs, this represents up to 70% of the normal human TCR vβ repertoire. **Figure 3A** shows representative flow cytometry plots of nine out of 24 TCR vβs measured. Vβ repertoire coverage for Mtb-specific IL-22-producing CD4+ T cells was broad and comparable to total CD4+ T cells, while slightly lower coverage was observed for Mtb-specific IFN-γ producing CD4+ T cells (medians 62%, 61% and 40%, respectively). Two TCRs, vβ2 and vβ5.1, accounted for more than 5% of the total vβ repertoire for both Mtb-specific IL-22 and IFN-γ producing CD4+ T cells (median: 13.6% and 10.4% for vβ2, 8.5% and 6.7% for vβ5.1, respectively), and were also the most prevalent vβs observed for total CD4+ T cells (median: 10.0% for vβ2 and 7.2% for vβ5.1, **Figure 3B**). Thus, TCR vβ repertoire usage was similar between IL-22 and IFN-γ-producing CD4+ T cells.

**Figure 3:**
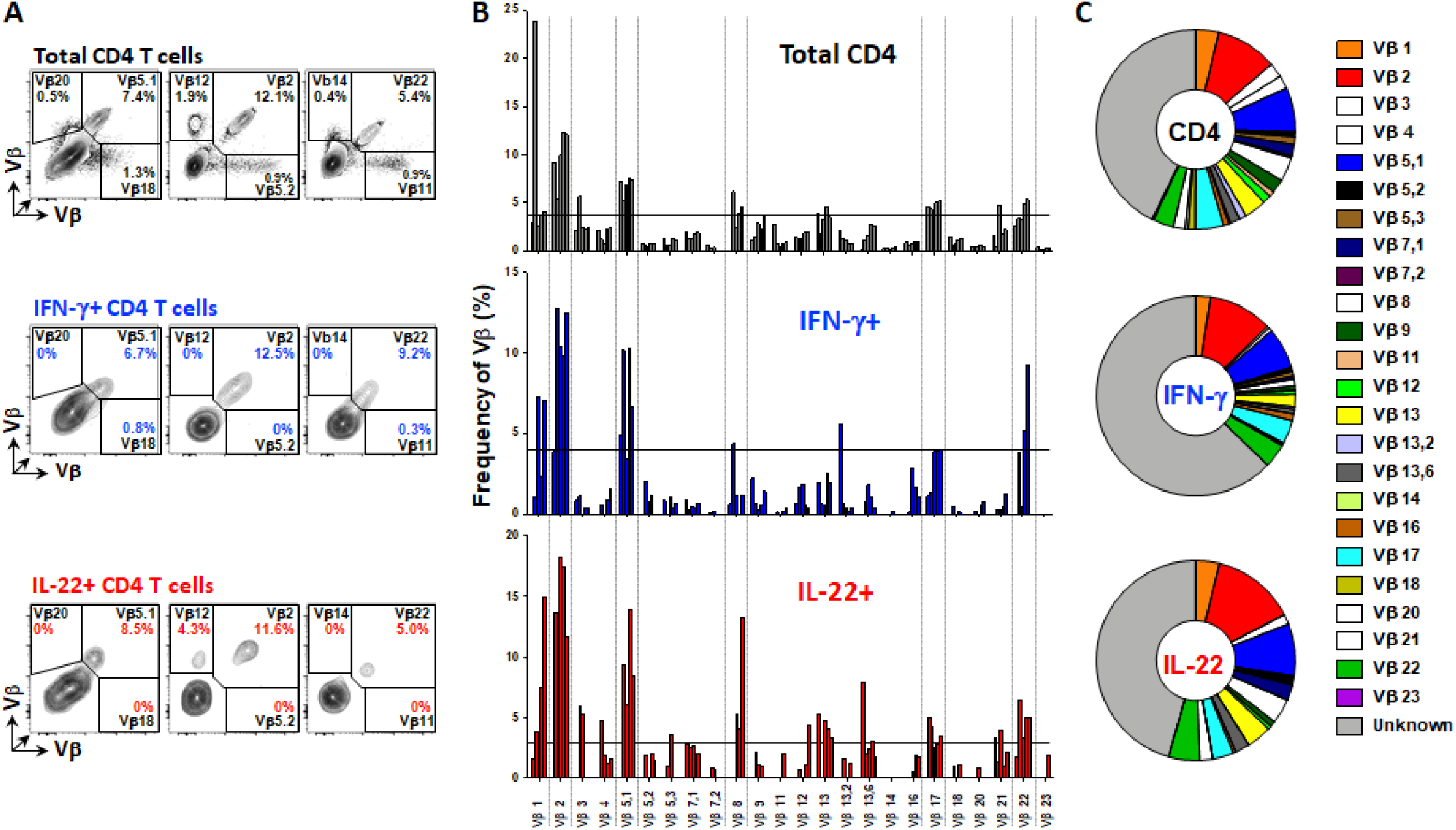
TCR Vβ repertoire of IFN-γ and IL-22-producing CD4+ T cells. **(A)** Representative flow cytometry plots showing staining of 9/24 Vβ receptors on total CD4+ T cells, IFN-γ and IL-22-producing CD4+ T cells. **(B)** Bar graph (each bar represent a donor) and **(C)** summary pie chart showing expression of 24 Vβ receptors by total CD4+ T cells, IFN-γ and IL-22-producing CD4+ T cells (n = 5). Means are depicted.

Overall, these results show that IL-22 contributes to a sizeable portion of the Mtb response and these cells constitute a subset distinct from Th1 or Th17 cells. Moreover, Mtb-specific IL-22-producing CD4+ T cells appear to be conventional CD4+ T cells and are dependent on TCR signaling for cytokine production.

### Mtb-specific Th22 cells exhibit distinct memory and activation profiles compared to Th1 and Th17 cells

To further describe the phenotypic characteristics of Mtb-specific Th22 cells and compare them to Th1 and Th17 subsets, we defined their memory (CD45RA, CD27 and CCR7) and activation profile (HLA-DR) in healthy individuals with latent Mtb infection. We focused our analysis on IFN-γ single producing cells (Th1), IL-22 single producing cells (Th22) and IL-17 single producing cells (Th17), since the proportion of cytokine co-expressing cells was negligible (**Figure 1C**). The measurement of CD45RA, CD27 and CCR7 enabled the detection of five distinct memory subsets, namely naïve (CD45RA+CD27+CCR7+), central memory (CM: CD45RA-CD27+CCR7+), transitional memory (TM: CD45RA-CD27+CCR7-), effector memory (EM: CD45RA-CD27-CCR7-), and effector cells (Eff: CD45RA+CD27-CCR7-; (**Figure 4A**). The memory profile of Mtb-specific CD4+ T cells varied depending on their Th polarization. Th1 cells were significantly enriched in the EM phenotype compared to Th22 and Th17 cells (median: 50%, 32% and 19%, respectively). Moreover, the Th22 subset was characterized by a low proportion of cells exhibiting a TM phenotype compared to the Th1 or Th17 subsets (median: 6%, 17%, 18%, respectively; **Figure 4C**).

**Figure 4:**
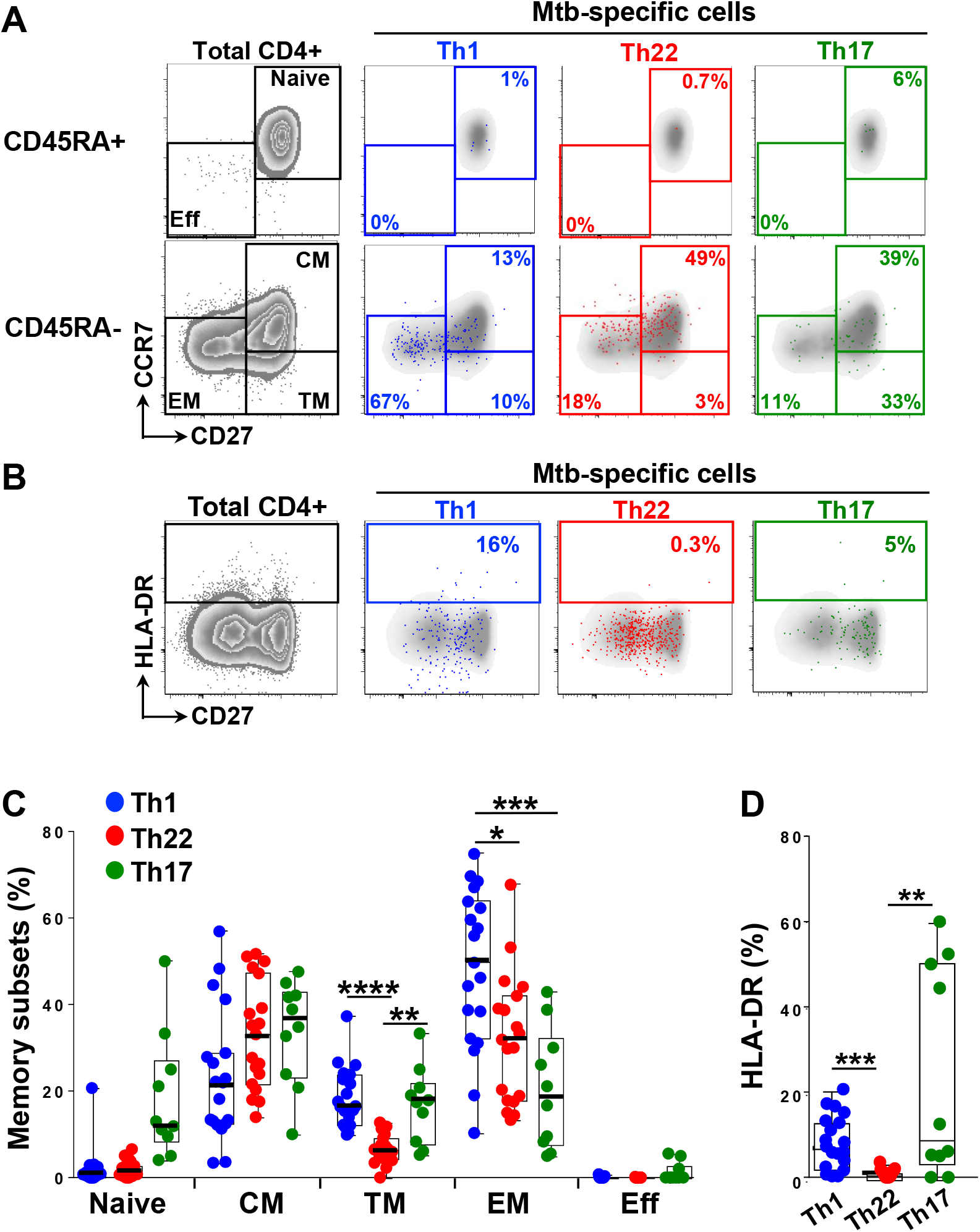
Comparison of the memory differentiation and activation profile of Mtb-specific Th1, Th22 and Th17 cells in LTBI individuals. **(A)** Representative overlay plots showing memory subsets in total CD4+ T cells (grey), IFN-γ+ (blue), IL-22+ (red) and IL-17+ (green) cells in response to Mtb lysate. (Naive: CD45RA+CD27+CCR7+; central memory (CM): CD45RA-CD27+CCR7+; transitional memory (TM): CD45RA-CD27+CCR7-, effector memory (EM): CD45RA-CD27-CCR7-and effector cells (Eff): CD45RA+CD27-CCR7-). **(B)** Representative overlay plots of HLA-DR expression in total CD4+ T cells (grey), IFN-γ+ (blue), IL-22+ (red) and IL-17+ (green) cells in response to Mtb lysate. Summary graph (n=19) showing distribution of memory subsets **(C)** and expression of HLA-DR **(D)** on IFN-γ, IL-22 and IL-17-producing CD4+ T cells in individuals with LTBI in the absence of HIV infection. Medians and interquartile ranges are depicted. Statistical comparisons were performed using a one-way ANOVA Kruskal–Wallis test. * *p* < 0.05, ** *p* < 0.01, *** *p* < 0.001, \*\*\*\**p* < 0.0001.

When assessing the activation status of Mtb-specific CD4+ Th subsets (**Figure 4B**), we observed that Th22 cells were characterized by a significantly lower expression of HLA-DR compared to both Th1 and Th17 cells (median 1.1%, compared to 7.3% and 9.3%, respectively; **Figure 4D**). The slightly increased frequency of HLA-DR-expressing Th22 cells observed during HIV infection, unlike Th1, merely mirrored background HIV-induced activation observed on total CD4+ T cells. It is plausible that other activation markers may be expressed on activated Th22 cells, and we have observed CD69 and CD25 upregulation on Th22 cells upon Mtb stimulation of blood from latently infected individuals (J. W. Milimu, R. Keeton, W. A. Burgers, unpublished).

Overall, these results indicate that Mtb-specific Th22 cells exhibit distinct memory and activation profiles compared to Th1 and Th17 cells.

### HIV infection and TB disease alter the distribution of Mtb-specific CD4+ Th subsets

To define the impact of HIV infection and TB disease on the distribution of Mtb-specific Th subsets, we compared the magnitude of Th1, Th22 and Th17 cells in 72 participants classified into four groups according to their HIV-1 and TB status: HIV-/LTBI (*n* = 19), HIV+/LTBI (*n* = 18), HIV-/aTB (*n* = 19), and HIV+/aTB (*n* = 16). The clinical characteristics of each group are summarized in **Table 1**. All participants were sampled prior to HIV and/or TB treatment. In participants with LTBI, HIV infection led to a significant reduction (median ∼3.6 fold) in the frequency of Mtb-specific Th1 cells (median: 0.23% for HIV+ and 0.84% for HIV-; p = 0.026; **Figure 5A**). While not statistically significant, the magnitude of the Mtb-specific Th22 response was also lower in HIV-infected participants compared to HIV-uninfected subjects (median: 0.18% and 0.45%, respectively). When assessing the effect of TB disease, we found that whilst TB disease did not significantly alter the magnitude of Th1 or Th22 responses in HIV-uninfected individuals, in HIV-infected persons the Mtb-specific Th profile was markedly distorted during TB compared to LTBI (**Figure 5A**). Indeed, the magnitude of Th1 responses was significantly higher in aTB/HIV+ individuals compared to LTBI/HIV+ participants (median: 1.21% vs 0.23%, respectively, p = 0.0005). In contrast, Mtb-specific Th22 cells were significantly lower in aTB/HIV+ compared to LTBI/HIV+ and LTBI/HIV-participants (median: 0.08% vs 0.23% and 0.45%, p = 0.046 and p < 0.0001, respectively). No differences were observed for Mtb-specific Th17 cells in all four clinical groups.

**Figure 5:**
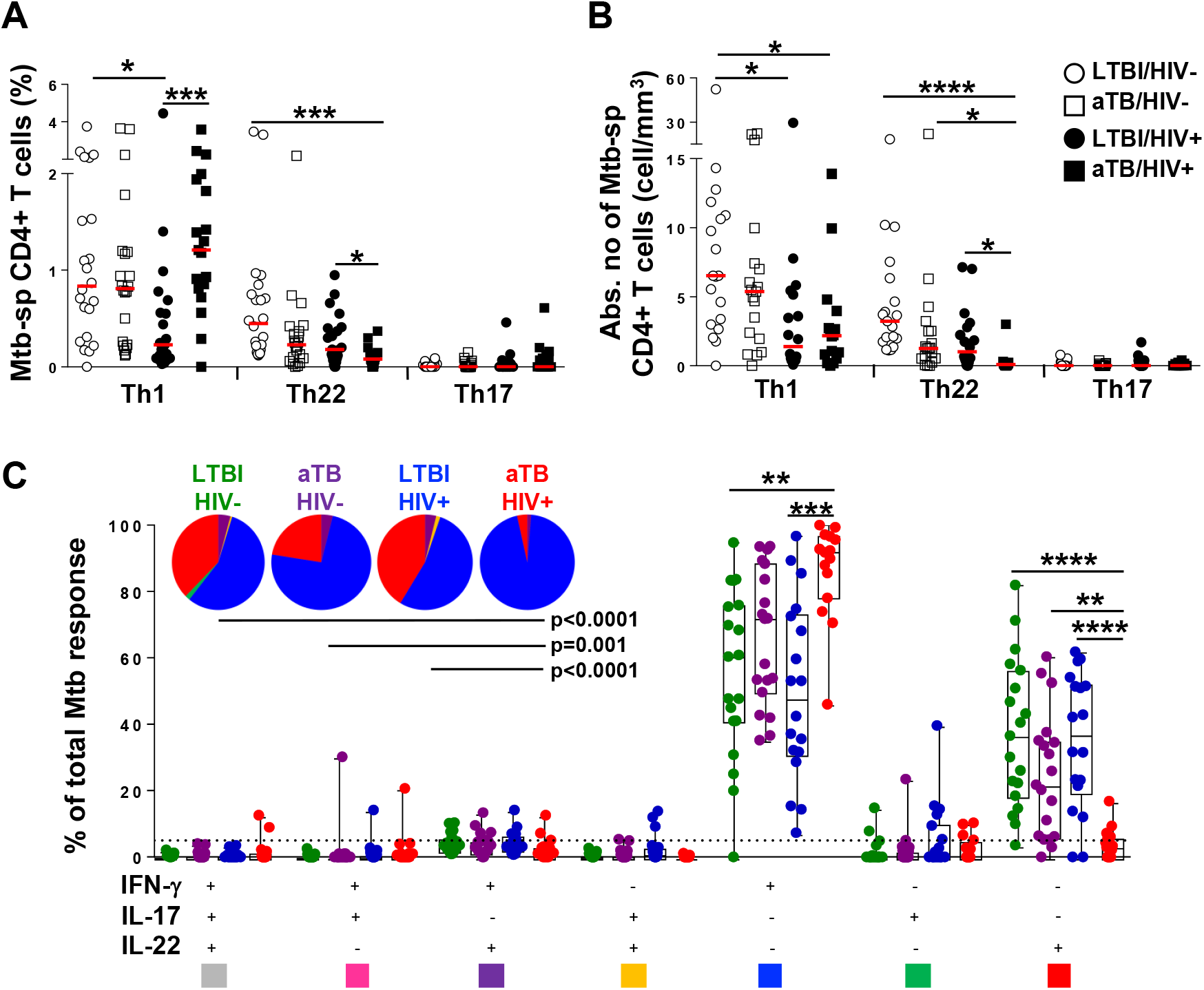
Contribution of IFN-γ, IL-22 and IL-17 to Mtb-specific CD4+ T cell responses and the effect of HIV infection and active TB disease. (**A)** Frequency of Th1, Th22 and Th17 responses and **(B)** absolute number of Th1, Th22 and Th17 cells detected in response to Mtb lysate in LTBI/HIV- (n = 19), aTB/HIV- (n = 19), LTBI/HIV+ (n = 18) and aTB/HIV+ (n = 16) individuals. Bars represent the medians. **(C)** Distribution of Mtb-specific cytokine responses. Each section of the pie chart represents median proportions of specific combinations of cytokines, as indicated by the color at the bottom of the graph. The bars represent the median and interquartile range. The dotted line is at 5%. Statistical comparisons were performed using a one-way ANOVA Kruskal–Wallis test. * *p* < 0.05, ** *p* < 0.01, *** *p* < 0.001, \*\*\*\**p* < 0.0001.

To account for the significant variation in absolute CD4+ T cell counts between groups (**Table 1**), the absolute number of Mtb-specific CD4+ T cells was calculated and compared between each group. As expected, HIV co-infection resulted in reduced Mtb-specific Th1 absolute cell numbers regardless of TB status (median: 1.4 vs 6.5 cells/mm^3^ in LTBI and 2.2 vs 5.4 cells/mm^3^ in aTB; **Figure 5B**). A comparable profile was observed for Th22 cells, but due to the decreased frequency of Th22 cells in aTB/HIV+ compared to LTBI/HIV+ and low CD4 counts in the former group, the absolute number of circulating Mtb-specific Th22 cells was markedly reduced in aTB/HIV+ individuals compared to LTBI (median: 0.09 vs 1.02 cells/mm^3^, respectively, **Figure 5B**). Of note, for HIV-infected individuals, we found no relationship between the frequency of any of the Th cytokine responses with absolute CD4 count or HIV viral load, regardless of TB status (data not shown).

Next, we examined how HIV infection and active TB might alter the relative contribution of Th subsets to the total Mtb response. Whilst there were no significant differences in the contribution of Th1 and Th22 cells to the Mtb response in LTBI/HIV-, LTBI/HIV+ and aTB/HIV-groups (medians for Th1: 60%, 48% and 72%; and medians for Th22: 37%, 37% and 22%, respectively), in the aTB/HIV+ group, the Mtb-specific response consisted almost exclusively of Th1 cells (>90%), with Th22 cells representing less than 5% of the total Mtb response (**Figure 5C**).

### TB disease and HIV co-infection differentially influence memory and activation profiles of Mtb-specific CD4+ Th subsets

Lastly, we performed phenotypic characterization of Mtb-specific CD4 Th subsets during HIV infection and TB disease. Comparing first the memory differentiation phenotype of Th subsets, we showed that TB disease, characterized by active bacterial replication, irrespective of HIV co-infection, promotes the differentiation of Mtb-specific Th1 cells. The proportion of Th1 cells exhibiting an EM phenotype was significantly higher in aTB compared to LTBI (69% vs 50% for HIV-, p = 0.0295 and 77% vs 45% for HIV+, p = 0.0004; **Figure 6A**). In contrast, no alteration of the memory differentiation phenotype of Th22 or Th17 cells was observed in aTB. We next assessed the effect of HIV infection and aTB on the activation profile of Mtb-specific CD4+ Th subsets. As previously described (36), irrespective of HIV infection, Mtb-specific Th1 cells in aTB were characterized by significantly higher expression of HLA-DR when compared to persons with LTBI (median: 45% vs 7%, respectively for HIV-, p = 0.0004 and 60% vs 8% for HIV+, p = 0.0009). However, aTB did not induce any significant changes in HLA-DR expression in Th22 or Th17 cells (**Figure 6B**).

**Figure 6:**
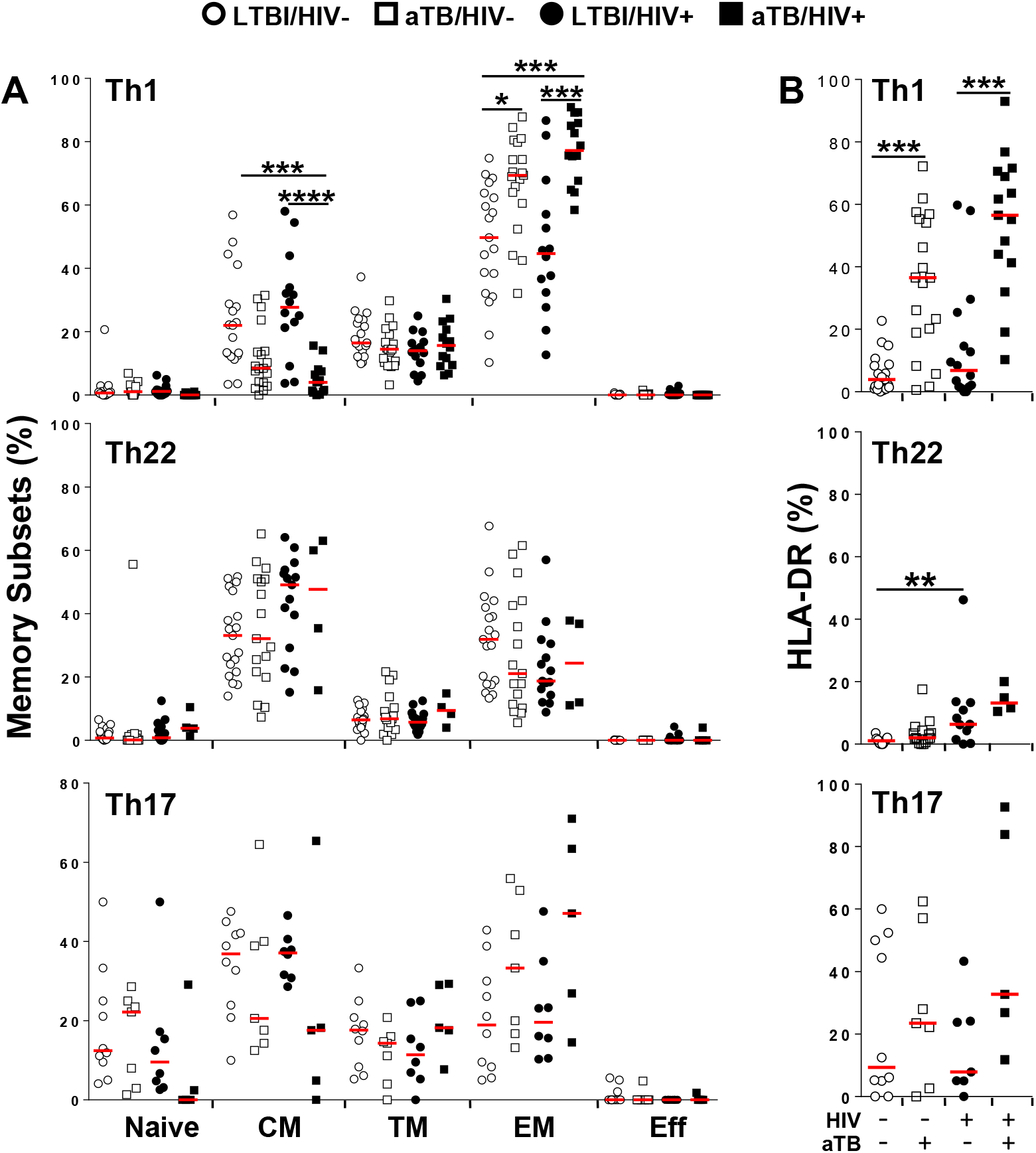
Memory and activation profile of Mtb-specific CD4+ T cells in individuals with distinct HIV and tuberculosis (TB) disease status. **(A)** Memory phenotype and **(B)** HLA-DR expression of IFN-γ, IL-22 and IL-17 producing CD4+ T cells in LTBI/HIV-, aTB/HIV-, LTBI/HIV+ and aTB/HIV+ groups. Red bars represent the medians. Statistical comparisons were performed using a one-way ANOVA Kruskal–Wallis test. * *p* < 0.05, ** *p* < 0.01, *** *p* < 0.001, \*\*\*\**p* < 0.0001.

These data confirm that Mtb replication during TB disease influences the memory differentiation and activation of Mtb-specific Th1 cells. In contrast, TB disease does not alter the memory or activation profile of Mtb-specific Th22 cells, indicating that TB differentially modulates Mtb-specific Th1 and Th22 cells.

## DISCUSSION

The importance of CD4+ Th1 (IFN-γ) and Th17 (IL-17) responses in protective immunity to *M. tuberculosis* is well established (9, 37). The emerging role of IL-22 and Th22 cells in TB immunity is less well studied, and we sought to address this knowledge gap. To better understand the contribution of Th22 cells to Mtb immune responses, we examined the dynamics of this subset in the context of TB disease and/or HIV infection by comparing the magnitude, differentiation and activation profiles of CD4+ T cells producing IFN-γ, IL-22 and IL-17. We confirm and extend our previous observations from latent Mtb infection (31), demonstrating that distinct, Mtb-specific IL-22-producing CD4+ T cells contribute a substantial portion to the total CD4+ T cell response to Mtb in both latent infection and TB disease. Furthermore, Th22 cells display different memory and activation profiles compared to Th1 and Th17 cells, and Mtb-specific Th22 are severely reduced in blood during TB disease in the context of HIV co-infection.

We show that Mtb-specific Th22 cells make up nearly 40% of the total Mtb response measured in both LTBI and in TB disease, with Th1 cells contributing 60% and Th17 cells less than 3% to the response. In some individuals, the Th22 response was 60-70% of the total CD4+ T cell response measured. These data suggest that Th22 cells may contribute to TB immunity. Indeed, IL-22 was recently found to play a protective role against the hypervirulent clinical strain of Mtb, HN878, in a mouse model (17). In this model, an indirect mechanism of Mtb control was reported, where IL-22 acted on lung epithelial cells to induce secretion of antimicrobial proteins, as well as the induction of chemokines that led to enhanced recruitment of macrophages (17). A direct effector function has also been described; whilst IL-22 receptors are present primarily on non-hematopoietic cells, there is mounting evidence that they may be expressed on macrophages (16, 17). IL-22 inhibited Mtb growth through induction of TNF-α to activate macrophages and enhance phagolysosomal fusion in infected macrophages through calgranulin A expression (16, 17, 38). In addition to these murine, non-human primate and *in vitro* models demonstrating a role for IL-22 in Mtb control, a human genetic study described polymorphisms in the promoter region of IL-22 leading to decreased IL-22 production, that were associated with greater TB susceptibility (26).

IL-22 has been classified as a Th17 cytokine due to co-expression with IL-17 in mouse studies and several shared functions between the two cytokines (39). Consistent with previous studies (29, 31), our results showed that the vast majority of IL-22 is produced independently of IL-17 (and IFN-γ) in humans. This implies a distinct role for IL-22 in the immune response to Mtb compared to IL-17 (40). Indeed, IL-22 was required for Mtb control at the chronic stage of infection, while IL-17 was important in the acute stage of Mtb infection in the same mouse model (17, 41). Although IL-22 and IL-17 have some overlapping functions, such as inducing antimicrobial peptides, regulating chemokine expression and promoting tissue proliferation and healing (17, 18, 39, 41), IL-22 may also mediate mycobacterial control by inducing TNF-α for macrophage activation (16, 17), a function that is more commonly associated with IFN-γ (42). Further studies are warranted to define the relative contribution and potential synergy of IL-22 with IFN-γ and IL-17 in protective immunity to TB.

In accordance with previous data (25), we show that Mtb-specific Th22 responses in the blood were 50% lower in individuals with TB compared to Mtb-exposed individuals. It is likely that in the context of TB disease, Mtb-specific Th22 cells migrate to the lungs. This conjecture is based on the fact that IL-22-producing cells have been detected in the lungs and granulomas of rhesus monkeys with TB (43, 44), that soluble IL-22 has been found to be elevated at the site of disease during both pulmonary and extra-pulmonary TB (24, 25), and also that IL-22-producing cells express CCR6, which has shown to mediate T cell homing to mucosal tissues (31). Two TB treatment studies further support this hypothesis; Suliman and colleagues found that patients with LTBI who received isoniazid prophylactic treatment had increased frequencies of BCG-specific IL-22-secreting CD4+ T cells compared to pre-treatment frequencies (45). Furthermore, Zhang and colleagues reported an increase in Mtb-specific soluble IL-22 in blood of TB patients at the completion of their anti-TB treatment, compared to before treatment (46). These studies suggest that reduction of antigen load at the site of disease led to recirculation of Th22 cells to blood.

Consistent with our recent findings (31), HIV infection resulted in a lower frequency of Th1 and Th22 cells in Mtb-exposed individuals, and also those with active TB. The depletion of Th22 cells could be explained by the fact that the majority of Th22 cells express CCR6 (31), and CD4+ CCR6+ cells display an increased permissiveness to HIV infection (47-49), are enriched in HIV DNA (50), and appear to preferentially support HIV replication (51). Indeed, CD4+CCR6+ cells have been shown to express high levels of the HIV co-receptors CCR5 and CXCR4, important for viral entry, as well as integrin α4β7, which has been associated with increased HIV susceptibility (47, 48, 52). Furthermore, CCR6+ cells lack the ability to secrete β–chemokines, which may protect against HIV in an autocrine manner (52). Thus, in the context of both TB disease and HIV infection, the combined effect of cell depletion and cell migration is the likely cause of the strikingly low IL-22 responses we observed in blood.

A range of innate and adaptive immune cells have been reported to produce IL-22 (16, 27, 26). In our study, we found that the predominant source of T cell-derived IL-22 in response to Mtb antigens was conventional CD4+ T cells, excluding MAIT, γδ and iNKT cells. Interestingly, whilst IL-22 responses were readily detectable upon stimulation with whole or lysed mycobacteria (*i*.*e*. Mtb cell lysate, gamma-irradiated Mtb and BCG), Mtb peptides were not able to stimulate IL-22 responses, raising the question of whether IL-22 production is induced through bystander effects on T cells from structural components present in these mycobacterial antigen preparations, thus leading to cytokine production. We showed for the first time that IL-22 production is indeed mediated by TCR engagement, as demonstrated by inhibition of TCR signaling and resultant abrogation of cytokine production. Furthermore, IL-22-producing CD4+ T cells displayed similar Vβ repertoire usage as IFN-γ producing CD4+ T cells. These data underscore our assertion that these are conventional CD4+ T cells; however, whilst, conventional, they may not be classically restricted (i.e dependent on MHC). The lack of detectable IL-22 in response to peptide stimulation suggests that induction of IL-22 may be via alternative antigen presentation mechanisms and/or additional co-stimulation requirements (53). One possibility is that IL-22 might be induced by modified peptide or non-peptide antigens, such as lipopeptides or lipids, and we are currently exploring this hypothesis. There is evidence supporting the production of IL-22 by CD4+ T cells recognizing CD1a (54, 55), and both CD1a and CD1c can present lipopeptide antigens to conventional T cells (56). Of note, group 1 CD1-restricted T cells specific for microbial lipid antigens display similar αβ TCR usage to peptide-specific T cells (57).

In conclusion, we provide evidence that Th22 cells are a major component of the specific adaptive immune response to Mtb during both infection and disease, and that these cells are reduced in blood during HIV co-infection, building on previous work. We hypothesise that Th22 cells could migrate to the lungs and expand for an effective secondary immune response to provide long-lasting protection against TB. To further investigate the specific role of Th22 cells in TB, it will be of interest to compare the profile of Th22 we observed in blood with the site of disease. The recent outcomes of vaccine candidate M72/AS01E demonstrating 50% efficacy against TB (5) and intravenous BCG vaccination showing sterilizing immunity in NHPs (7) provide an ideal setting to assess the potential protective role of Mtb-specific Th22 cells.

## Data Availability

All data generated or analysed during this study are available from the corresponding author (WAB) on reasonable request.

## Data Availability

All the reported data are available within the manuscript

## ACKNOWLEDGMENTS

We thank the study participants for providing samples and for their time and commitment to the study, and to the clinic staff at the Ubuntu HIV-TB clinic. We thank Dr Charlotte Schutz and Dr Cari Stek for phlebotomy. We thank Tracey Müller for assistance with sample processing, and Kathryn Norman for administrative assistance. The following reagents were obtained through BEI Resources, NIAID, NIH: *Mycobacterium tuberculosis*, Strain H37Rv, Gamma-Irradiated Whole Cells, NR-14819 and *Mycobacterium tuberculosis*, Strain H37Rv, Whole Cell Lysate, NR-14822.

## CONTRIBUTIONS

MSM, RJW, CR and WAB designed the study; MSM, RB and FMAO performed the experiments; MSM analyzed the data and wrote the paper. CR and WAB contributed to the data discussion and writing the paper. All authors read and approved the final manuscript.

## Footnotes

This project is part of the EDCTP2 programme supported by the European Union (EU)’s Horizon 2020 programme (TMA2016SF-1535-CaTCH-22, to WAB). WAB was also funded by the SAMRC, NRF SA (92755) and NHLS Trust (2016-2DEV04). Further funding came from the National Institutes of Health, the Office of the Director (OD) (NIH, R21AI115977 to CR). CR is supported by the EDCTP programme supported by the European Union’s Horizon 2020 programme (TMA2017SF-1951-TB-Spec, to CR). RJW is supported by the Wellcome Trust (203135 and 104803), NIH (U01 AI115940), the Francis Crick Institute (Cancer Research UK, MRC UK and Wellcome FC0010218), and SAMRC (SHIP). The funders had no role in study design, data collection and analysis, decision to publish, or preparation of the manuscript.

---

**Supplemental Figure S1:**
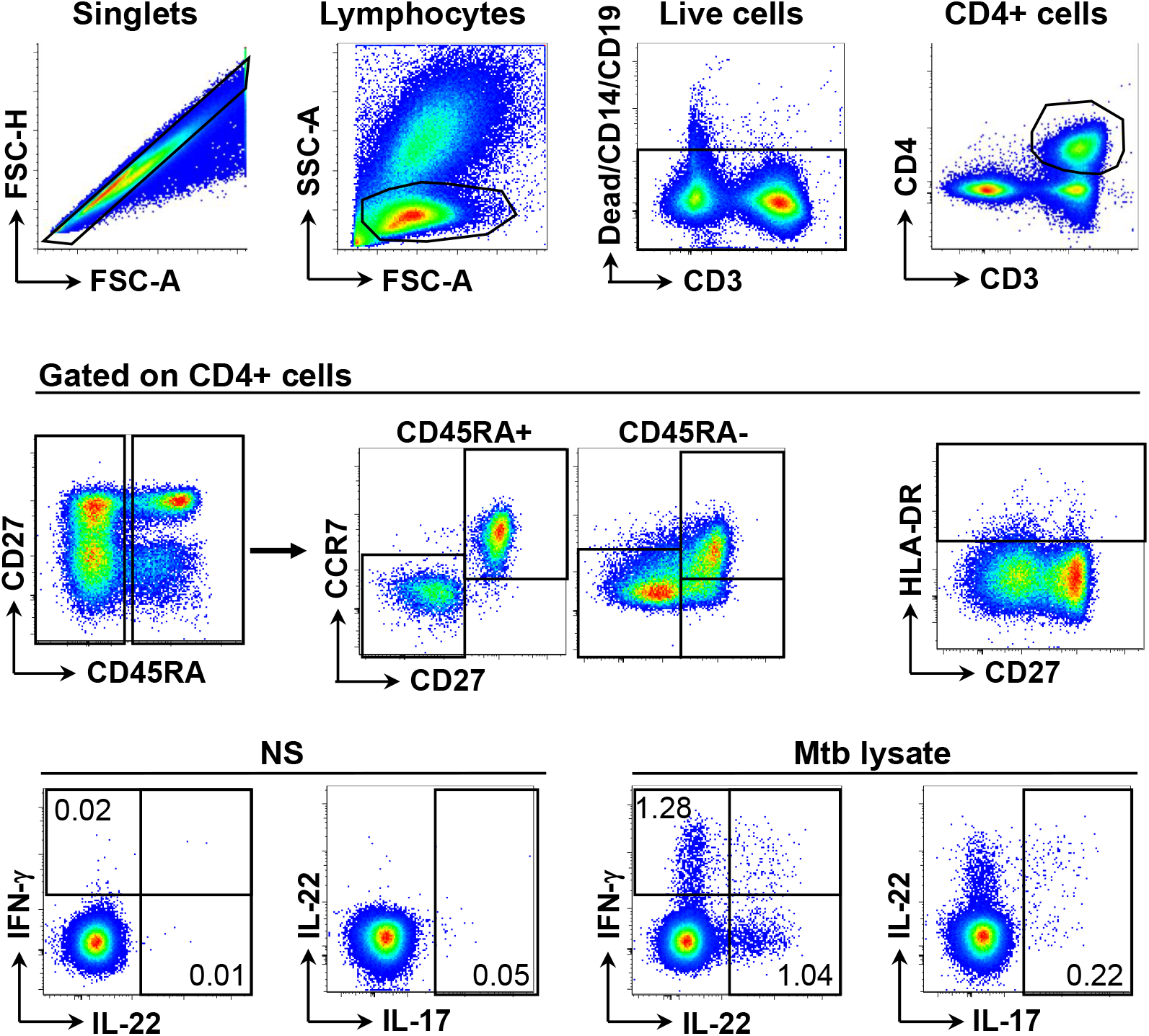
Gating strategy. Gating strategy used to determine the phenotype, memory profile and functional properties of CD4+ T cells. NS: non-stimulated.

**Supplemental Figure S2:**
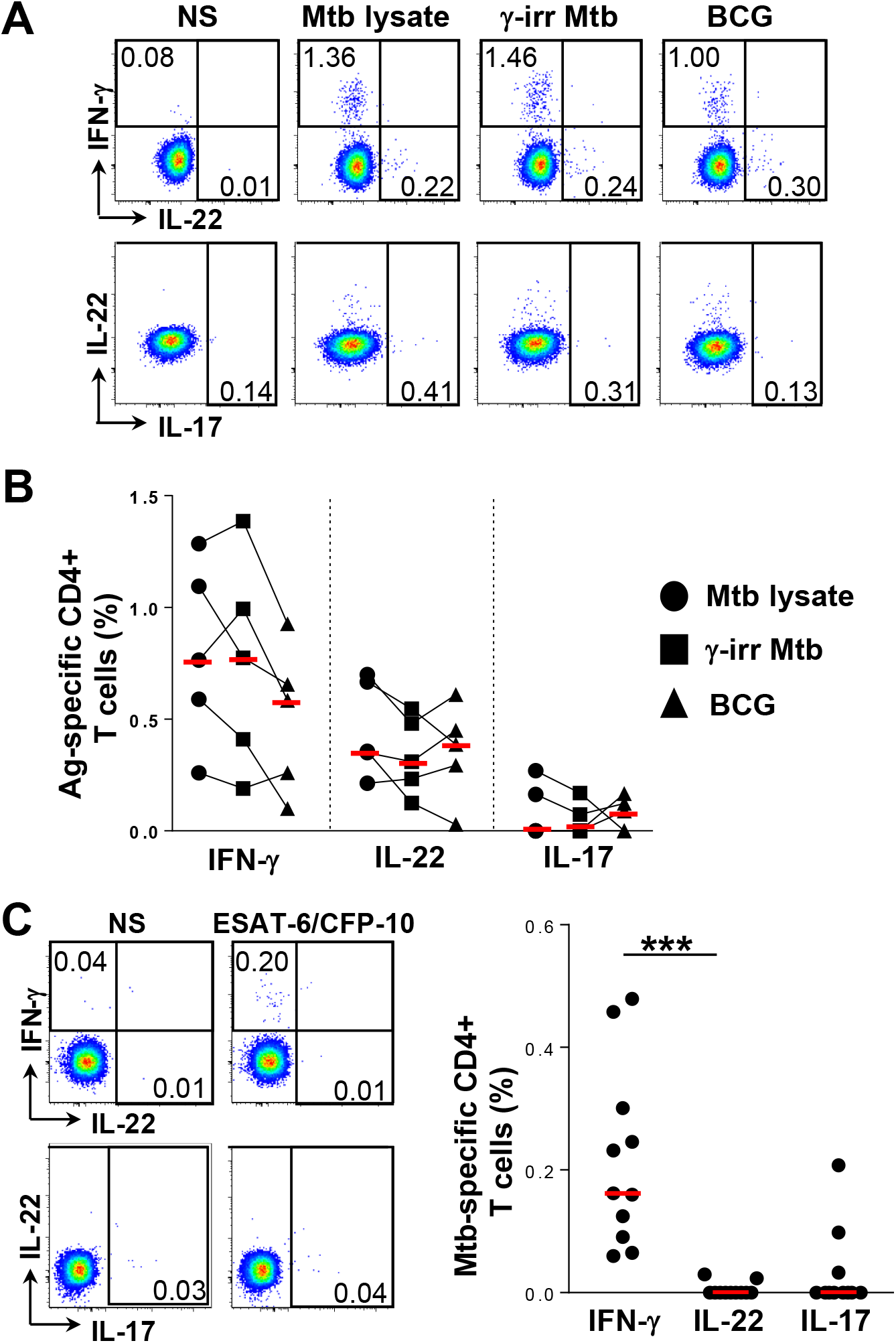
Comparison of CD4+ T cell frequencies detected using Mtb lysate, gamma-irradiated Mtb, and BCG. **(A)** Representative flow cytometry plots showing IFN-γ, IL-22 and IL-17 responses after stimulation of blood from healthy donors with Mtb lysate, gamma-irradiated Mtb, and BCG. **(B)** Comparison of the frequency of IFN-γ, IL-22 and IL-17 after stimulation with different antigens (n=5). **(C)** Comparison of the frequency of IFN-γ, IL-22 and IL-17 when stimulated with ESAT-6/CFP-10 (n=11). Red bars represent the medians. Statistical comparisons were performed using a one-way ANOVA Friedman test. *** *p* < 0.001.

